# A mHealth intervention (mTB-Tobacco) for smoking cessation in people with drug-sensitive pulmonary tuberculosis: protocol for an adaptive design, cluster randomised controlled trial (Quit4TB)

**DOI:** 10.1101/2024.12.10.24318761

**Authors:** Maham Zahid, Fahmidur Rahman, Ai Keow Lim, Asiful Chowdhury, Shakhawat Hossain Rana, Saeed Ansari, Mahmoud Danaee, Melanie Boeckmann, Steve Parrott, John Norrie, Amina Khan, Rumana Huque, Kamran Siddiqi, the RESPIRE collaboration

## Abstract

**Introduction:** People with tuberculosis (TB) who continue to smoke are more likely to have poor health outcomes than those who quit. Established smoking cessation approaches such as mHealth may help TB patients quit smoking. This paper summarises the methodology proposed to assess the effectiveness and cost-effectiveness of mTB-Tobacco (an mHealth intervention) in helping TB patients stop smoking and have improved health outcomes.

**Methods and analysis:** A two-arm, parallel, open-label, multi-centre, cluster randomised, two-stage adaptive design trial is proposed to first evaluate the superiority of mTB-Tobacco, compared with usual care and then the non-inferiority of mTB-tobacco compared with face-to-face behaviour support. Study settings include TB treatment centres in Bangladesh and Pakistan. The study population includes adult patients, newly diagnosed (within four weeks) with pulmonary TB disease, daily smokers, willing to quit, and have access to mobile phones. The primary outcome includes biochemically verified continuous smoking abstinence assessed at 6 months per Russell Standard. A generalised linear mixed-effects model will be used to assess the impact of mTB-Tobacco intervention on continuous outcomes, incorporating fixed effects for the intervention, random effects for clusters, and relevant covariates. Cost-effectiveness analysis will be done to estimate the cost per quitter and cost per QALY gained, calculate the incremental cost-effectiveness ratios (ICER) to establish the value for money for mTB-Tobacco.

**Ethics and dissemination:** This trial will be conducted in compliance with ICH-GCP guidelines and the Declaration of Helsinki. The study has been approved by the ethics committees of the University of Edinburgh Medical School Research Ethics Committee (EMREC) of UK, the Bangladesh Medical Research Council (BMRC) and the National Bioethics Committee (PMRC) of Pakistan.

**Funding:** The study is funded by the National Institute of Health Research (NIHR) UK under a research award named NIHR Global Health Research Unit on Respiratory Health (RESPIRE) (Award ID: NIHR132826)

**Trial registration number:** ISRCTN86971818 (https://doi.org/10.1186/ISRCTN86971818); Submission date:29/08/2023; Registration date:11/09/2023; Last edited:30/04/2024

**Strengths and limitations of this study:**

- This trial will be the first one to test the effectiveness of mHealth based intervention to help TB patients quit smoking at 6 months.
- An effective mTB-Tobacco intervention could be transformational for TB patients who smoke. It will not only benefit the TB patients but will also allow the national TB control programmes to have a less resource-intensive and effective intervention for smoking cessation that can be easily integrated into their system.
- The web portal that will be developed under this project can be used by TB programmes with no to minimal extra cost to only account for the SMS text message delivery.
- TB disease is more prevalent in vulnerable populations e.g. lower socioeconomic groups, and such population is more likely to have poor literacy as compared to the general population. This can be a challenge as TB patients’ ability to read and understand SMS messages may be a barrier.

## INTRODUCTION

Tuberculosis (TB) is an infectious disease with high morbidity, mortality and economic burden. Despite being preventable and curable, TB remains the world’s second leading cause of death from a single infectious agent in 2022, the first being coronavirus disease (COVID-19). The Global Tuberculosis Report 2023 states that in 2022, there were 7.5 million reports of newly diagnosed TB, which is highest in the history perhaps due to diagnosis and treatment delays faced during COVID-19 pandemic, followed by 1.3 million deaths.^1,2^ The majority of the TB burden remains concentrated in low and middle-income countries (LMIC), including Pakistan and Bangladesh. A downward trend of TB burden has been noticed in all World Health Organisation (WHO) regions, but the decline is not sufficient to end the global TB epidemic in the context of Sustainable Development Goals (SDGs).^3^

Amongst five major attributable risk factors including malnutrition, HIV infection, smoking, alcohol use and diabetes, 0.7 million new TB cases are attributable to smoking alone, especially in males.^4^ A meta-analysis conducted in 2022 to assess the effect of smoking on TB treatment outcome concluded that smoking increases the chances of poor treatment outcome by 51% (OR=1.51, 95% CI 1.30-1.75, I-square 75.1%), where the effect was even higher for LMIC (OR=1.74; 95% CI 1.31-2.30).^5^ Another meta-analysis concluded that smokers had greater odds of unfavourable outcomes (pooled OR=1.23, 95% CI 1.14–1.33), delayed sputum conversion (pooled OR=1.55, 95%CI 1.04–2.07), and loss to follow-up (pooled OR=1.35, 95%CI 1.21–1.50).^6^

In terms of tobacco use, 1.3 billion people globally use tobacco in one or more forms, and 80% of them live in LMIC.^7^ A dual burden of TB and tobacco use in these countries makes it imperative to devise strategies that address both of these conditions. Literature shows that there are significant benefits of quitting smoking on TB treatment outcomes. Those who quit smoking may have better TB cure rates (91% vs 80%, p<0.001), lower TB relapse rates (6% vs 14%, p<0.001), and higher sputum smear conversion at 2-months (91% vs 87%, p=0.036), than as compared to those who continue did not quit smoking, respectively.^8^ This makes smoking cessation in TB patients a public health priority. There is a need for evidence-based smoking cessation approaches^9^ and increasing policy support to help TB patients quit smoking.^10^

Smoking prevalence in TB patients is higher than the general population,^11^ but the vast majority of TB patients are neither routinely asked about their smoking status nor advised to quit.^12^ There are well-established pharmacological and non-pharmacological interventions developed and tested for effectiveness to help TB patients quit smoking. Previous randomised controlled trials (RCT) in Bangladesh and Pakistan have shown that face-to-face behavioural interventions are effective for smoking cessation, reporting 41% quit rate.^13^ Despite recognising the benefit of quitting and ownership by policymakers, the integration of the smoking cessation package in routine TB care remains challenging. Due to health system barriers including insufficient human resources, cost, reach and sustainability^14^ no TB high-burden countries have so far integrated face-to-face smoking cessation within TB services.

Recognising the challenges of integrating and scaling up face-to-face interventions, the WHO developed a mHealth smoking cessation package (mTB-Tobacco) that can be delivered as SMS (Short Message Service) messages via mobile phones to TB patients.^15^ It is anticipated that delivering mHealth package will cost less than resource-intensive face-to-face behavioural interventions, but it is unknown if this package is as effective and therefore more cost-effective than face-to-face behavioural intervention. Hence, there is a need to first establish the effectiveness of mHealth intervention in terms of helping TB patients to quit smoking. The Quit4TB trial is designed to assess the effectiveness and cost-effectiveness of mTB-Tobacco in achieving continuous abstinence for at least six months and improving health outcomes in people with TB who smoke daily.

## METHODS AND ANALYSIS

### Trial design

The Quit4TB trial is a two-stage adaptive design, multi-centre, cluster randomised controlled trial consisting of four phases as shown in figure 1:

**Figure 1:**
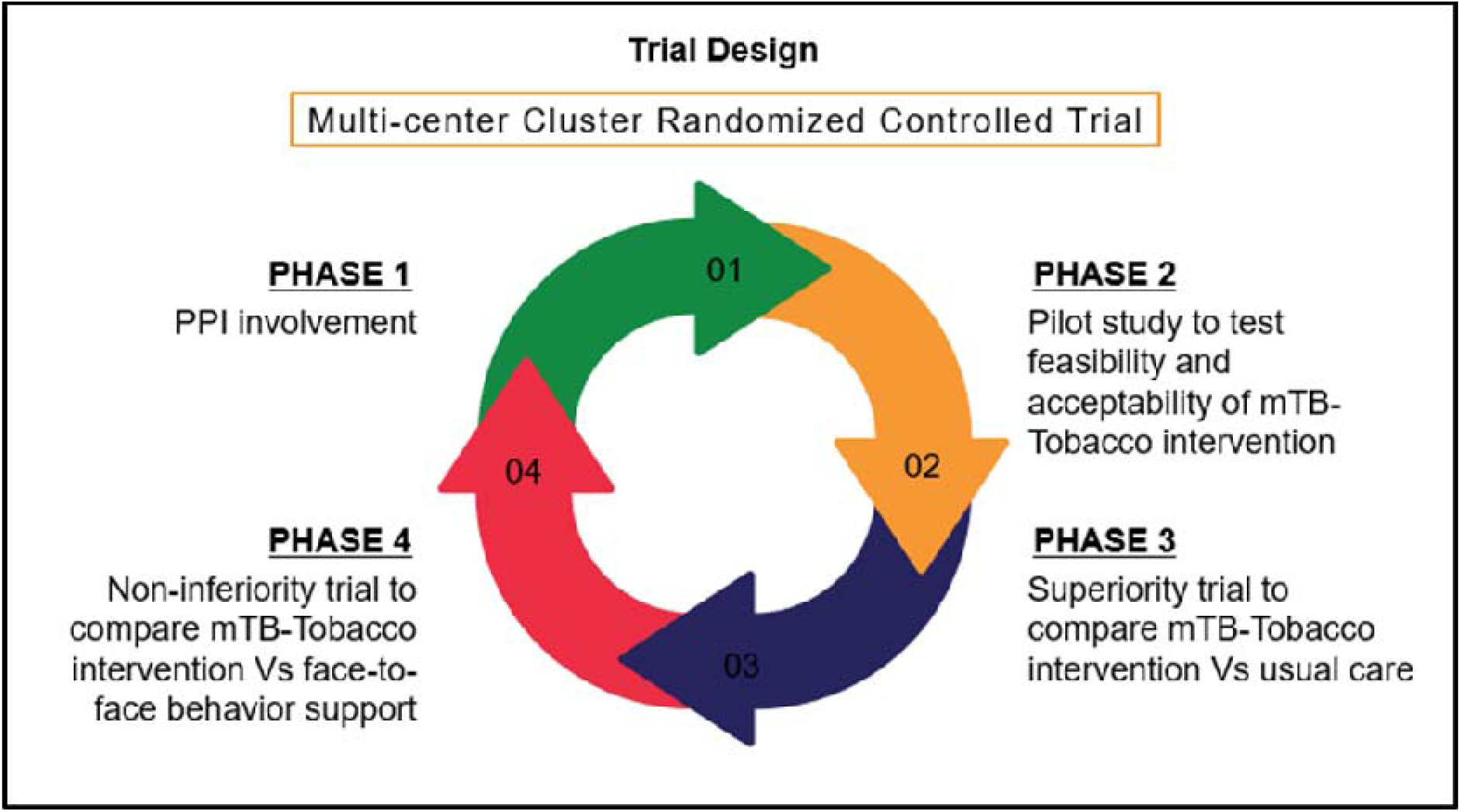
Study design and phases of Quit4TB trial.

**Phase 1** involves consultations with a Patients and Public Involvement (PPI) group about the study processes. PPI members will review and provide feedback on the SMS text messages translated into local languages as well as the participant information sheet and consent forms. For this, a PPI group of 7-8 members will be formed including TB patients, their family members, health care professionals and public representatives, and translated study materials will be discussed with them individually or as a group. Debriefing will be done after PPI engagement, and necessary adjustments will be made based on the feedback.

**Phase 2** consists of a pilot study, which aims to assess the feasibility of the mTB-Tobacco intervention and other trial processes. The mTB-Tobacco web-based intervention will be pilot –tested with a total of 16 participants, 8 each from Bangladesh and Pakistan, where equal half of the participants will belong to the intervention and control groups. The pilot data will be evaluated using two sets of data, users’ self-reported experiences and their real-time engagement with the programme. Users’ experiences will be collected at a 9-week follow-up through face-to-face interactions. Key questions will focus on users’ experience of taking part in the mTB-Tobacco programme; the clarity, quantity, timing and frequency of messages; what was good about the programme and what was not; completion or non-completion of the programme; and any effect on their attitudes or target behaviour(s). Users’ real-time engagement will be evaluated using computer records from the programme used for launching mTB-Tobacco. The pilot study will be embedded in the next phase of the study, by continuing the pilot study participant’s follow-up up to 6 months, and will be treated with the same processes and governance as the main trial participant data.

**Phase 3** will be the superiority trial, which will last for 12 months (6 months recruitments and 6 months follow-ups), to compare mTB-Tobacco (intervention A) with usual care (control).

**Phase 4** will be the non-inferiority trial that will be conducted if in phase 3 the mTB-Tobacco intervention turns out to be effective in helping TB patients quit smoking as compared to control. Phase 4 will then last for another 12 months (6 months recruitment and 6 months follow-ups), to compare mTB-Tobacco (intervention A) with the face-to-face behavioural support (intervention B) to assess if it’s as effective as the face-to-face behavioural support intervention to help TB patients quit smoking.

The trial flow chart is given in Figure 2, followed by a written explanation of various aspects of the trial.

**Figure 2:**
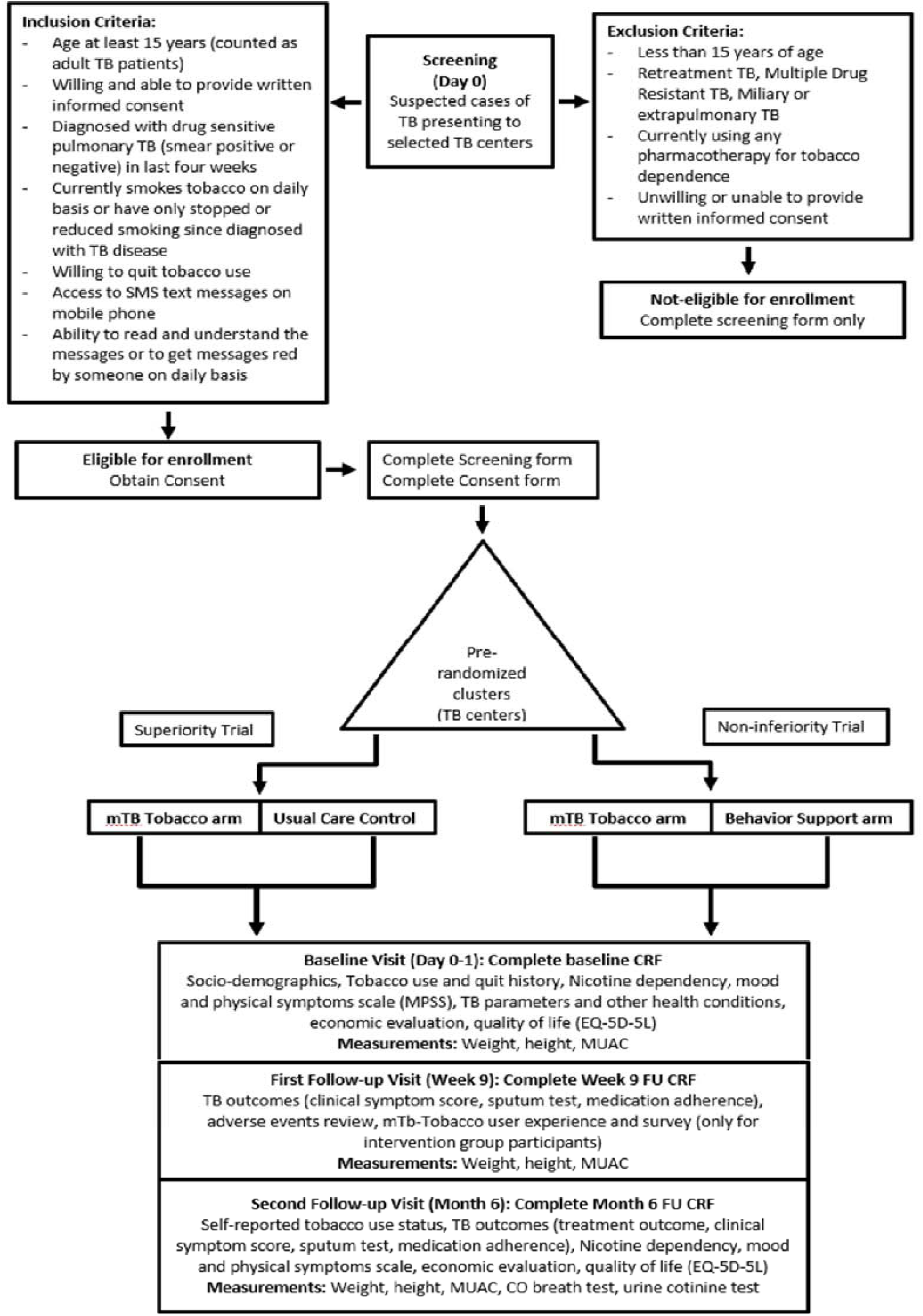
Quit4TB trial flow chart.

### Study settings

This trial will be conducted in Bangladesh (Dhaka division) and Pakistan (Punjab province) simultaneously. A prespecified number of clusters (TB centres) will be selected from each country to recruit study participants.

### Eligibility criteria for inclusion of clusters

All TB centres with a minimum turnover of 50 new TB patients per month will be eligible for selection, to make sure that adequate participant enrolment target is achieved. The included sites must be functioning and designated TB diagnostic centres approved by the National /Provincial TB Control Programme (NTP/PTP). These may include primary, secondary, or tertiary care health centres (HCs), generally known as the District/or Tehsil headquarter hospitals in Pakistan. In Bangladesh, health facilities will be the Directly Observed Therapy (DOTs) corners at Upazila (sub-district) Health Complex and Urban DOTS corners. In addition, the facility must be able to nominate a site focal person (an NTP personnel e.g., the district TB coordinator, or the responsible TB clinician at the site) who assumes responsibility for the proper conduct of the trial at their site; the health facility should have an adequate number of qualified staff and adequate facilities for the foreseen duration of the trial to conduct the trial properly and safely, and the HC should be willing to join the trial and provide required administrative support. Facilities not meeting the inclusion criteria will not be eligible for the trial.

### Study participants

TB patients presenting at selected TB centres (clusters) will be screened for inclusion in the trial as per the following eligibility criteria:

### Eligibility criteria for inclusion of participants

Participants aged 15 years or older, diagnosed with drug-sensitive pulmonary TB (smear positive or negative) in the last four weeks, currently smoking tobacco daily or have only stopped or reduced smoking (less than daily) since diagnosed with TB, willing to quit tobacco use, have access to personal mobile phone and willing and able to provide written informed consent will be included in the study. Informed consent will be taken from adult participants (18 years and above), while assent along with parent/guardian’s consent will be taken for adolescent participants (15-17 years of age). Exclusion criteria include patients anticipated to have adverse effects either due to the study treatment or research burden. These will include those who are/have less than 15 years of age, retreatment TB, MDR TB, miliary or extrapulmonary TB, currently using any pharmacotherapy for tobacco dependence and unwilling or unable to provide written informed consent.

### Randomisation

Since the interventions will be delivered at the group level, this trial is designed as a cluster randomised controlled trial (cRCT) where TB centres will be randomly allocated to the different trial arms. The TB centres (clusters) would be randomised in 2:2:1 ratio to mTB-Tobacco (intervention A), face-to-face (intervention B), or standard care (Control). An independent statistician, blinded to clusters, will identify and use computer-generated random-number lists to generate the allocation sequence. The allocation will be based on using minimisation to ensure balance across the groups on the average number of TB patients seen per month and geographical locations (Bangladesh and Pakistan).

### Blinding

In this study, we utilised a blinding methodology to guarantee that specific key individuals remained uninformed about the allocation of treatments. The allocation sequence will be determined by an independent statistician, who has no knowledge of the participating facilities. Nevertheless, as the SMS receipt is a noticeable component, participants will be able to determine whether they are receiving the SMS or not. Therefore, it is not possible to achieve total blindness at the participant level in this circumstance. In addition, the principal investigator (PI) and the research teams are aware of the specific cluster assigned to each treatment group. However, steps will be taken to potentially prevent the data analyst from knowing the treatment assignments in order to reduce bias during the analysis of the data. Thus, the blinding in our study mostly happened with the independent statistician who randomly assigned the clusters to interventions, and the data analyst as well.

### Treatment and control arms

#### 1. mHealth based mTB-Tobacco intervention

The mTB-Tobacco is an mHealth intervention, based on a smoking cessation package developed by the WHO that can be delivered as mobile SMS text messages to TB patients.^17^ This package comprises two distinct areas of intervention, the first tries to instil behaviour change in patients to quit the habit of tobacco use and the second includes a combination of TB-related supportive motivational and informative messages. The package aims not only to inform TB patients about the hazards of tobacco use and encourage them to quit, but also to enhance treatment adherence, increase healthy behaviours, and reduce potentially harmful behaviours in TB patients to improve their overall health outcomes.

The set of SMS text messages included in the mTB-Tobacco package was adapted by research experts to include 134 one-way messages, and a transmission schedule was designed to set the days for each message delivery along with a specified time. Each participant in the intervention group will receive a total of 134 SMS messages over 6 months. In the first two months, the frequency of messages would be 4 to 5 messages per day, in the next two months the frequency would reduce to 1 to 2 messages per day and in the last two months there will be 1 message sent after a couple of weeks. The final list of messages and delivery schedule is given as Supplementary Table I.

The messages will be translated into local languages including Urdu and Bangla by local research teams, using the translation and validation steps recommended by the WHO.^16^ The initial forward translation will be carried out by an independent bilingual translator in local language, the translated content was back-translated into the English language by another independent translator. The conceptual and cultural context will be taken into account. The discrepancies will be discussed with the local principal investigators and research team to make translations satisfactory.

A web-based digital application (mTB-Tobacco) will be developed by the local research team to sync the data with the database and send the SMS text messages automatically to the enrolled participant’s personal mobile numbers, following the set schedule. The mTB-Tobacco web portal will store the participant’s unique ID number and active mobile number for the trial. No other personal or identifiable information will be stored.

#### 2. Face-2-face behavioural support (Intervention B)

This consists of an adapted version of a pre-developed and proven effective intervention to help people quit smoking and smokeless tobacco use.^9^ This consists of two face-to-face sessions delivered on day 0 and day 5 (+2) which last 10 and 5 minutes respectively. The sessions will be structured using an educational flipbook; the session on day 0 will be aimed at encouraging tobacco users to see themselves as non-users and set a plan for a quit date five days later; with a session on the quit date (day 5) to review progress. Further encouragement and support (if needed) will be offered at a subsequent visit in week 5.

#### 3. Standardised usual care (Control)

In the usual care group, the patients will undergo routine TB treatment care. In addition, education leaflets will be given to the control arm participants. The leaflets containing information on the harmful effects of tobacco and advice on stopping smoking will also be given to the participants belonging to the intervention arms A and B.

### Study Outcomes

#### Primary outcomes

The primary outcome will be biochemically verified continuous abstinence at 6 months post-randomisation.^18^ Abstinence is defined as a self-report of not having used more than 5 cigarettes, bidis, water pipe sessions since the quit date, verified biochemically by a breath carbon monoxide (CO) reading of less than 10 ppm at month 6.^18^

In case of concomitant smokeless tobacco use, biochemical verification will be carried out using COT Rapid Test Cassette (Hangzhou All Test Biotech Co. Ltd.) to detect cotinine (a nicotine metabolite) level in urine samples. The test result will be interpreted as “negative”, if two lines appear (one coloured line in the control line region, and another apparent coloured line in the test line region) or “positive”, if one coloured line appears in the control line region. The negative result will indicate that the cotinine concentration is below the detectable level (200 ng/mL) while the positive result will indicate that the cotinine concentration exceeds the detectable level (200 ng/mL). The RA will perform cotinine dipstick tests on urine samples collected from the patients, which will be discarded after cotinine levels have been recorded on the CRFs.

When a patient self-reports abstinence with an elevated CO level of >10ppm (and cotinine levels in concomitant users indicating active tobacco use), the biochemical verification will supersede the self-report and the patient will be defined as a tobacco user.

#### Secondary outcomes

The secondary outcomes include: Point abstinence, defined as a self-report of not using tobacco in the previous 7 days, assessed at week 9 and month 6; Adherence to TB Treatment: All registered TB patients’ medication logs (for anti-TB medication) are recorded on the ‘Treatment Support Card’; a copy of this card would be requested from the TB paramedic by the RA and attached with the patient CRF; TB Programme outcomes: the proportion of treatment success (including cured and completed treatment), treatment failure, defaulted and died will be recorded from TB register (TB03) at month 6. Table 1 gives definitions of TB treatment outcomes.

**Table 1:** Definitions of TB treatment outcomes as per guidelines.

| **Sr. no.** | **TB Treatment Outcomes** | **Definition** |
| --- | --- | --- |
| **1.** | Cured | A patient who was initially smear-positive and who was smear-negative in the last month of treatment (at month 6) and on at least one previous occasion |
| **2.** | Completed treatment | A patient who completed treatment (at month 6) but did not meet the criteria for cure or failure |
| **3.** | Treatment failure | A patient who was initially smear-positive and who remained smear-positive at month 6 or later during treatment |
| **4.** | Defaulted | A patient whose treatment was interrupted for 2 consecutive months or more |
| **5.** | Died | A patient who died from any cause during treatment |
| **6.** | Relapse | A patient who was previously treated for TB, was declared cured or treatment completed at the end of his most recent course of treatment and is now diagnosed with a recurrent episode of TB (either a true relapse or a new episode of TB caused by reinfection). |

### Study assessments

All study participants will have assessments on day 0, week 9 and month 6 time points corresponding with routine TB clinic visits. All the trial activities will be performed by field research assistants (one RA per site) who will be hired and trained to take patient consent, collect baseline data, follow up data and perform study assessments. The trial assessment schedule (table 2) outlines the visit dates necessary for data collection, but the patient may be seen more frequently for routine clinical care as needed. During the treatment period, if study participants are unable to attend on the day, every effort would be made to complete the visit within 2 days of the scheduled date. If a scheduled visit is missed without notice, then the RA should endeavour to contact the patient by phone or by home visit.

**Table 2:**
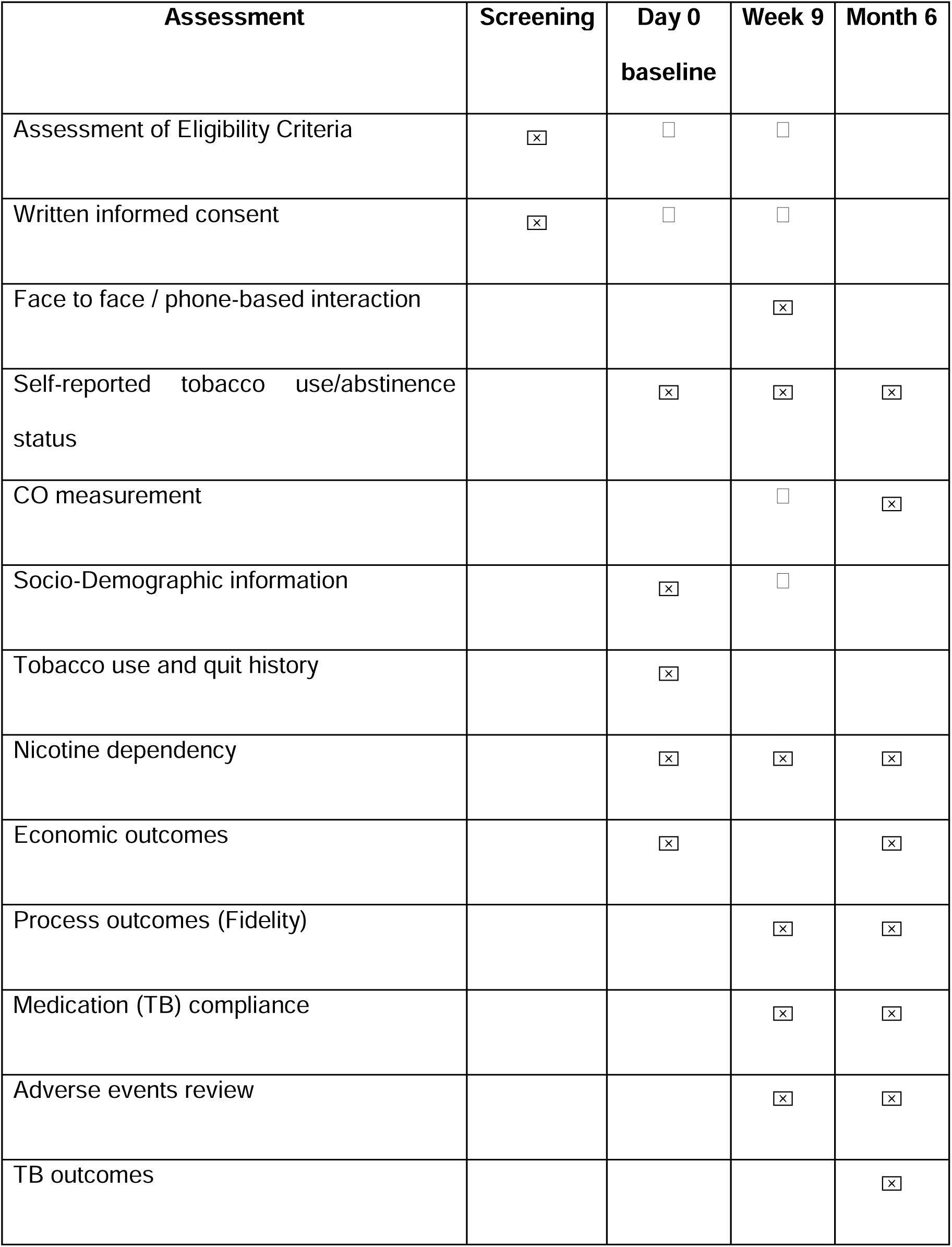
Participant enrolment and follow-up assessment schedule for Quit4TB trial.

During the follow-up phase, scheduled assessments will be carried out no more than 5 days before the scheduled visit. Scheduled visits will be rescheduled to allow for public holidays or other unavoidable circumstances that affect the scheduled visit date, but the re-scheduled visit should be no more than 5 days from the originally scheduled visit date. Participants will also be given a card with the contact details for the trial RA and the clinical TB care team at their site. If a patient is more than 5 days late for a scheduled study visit, an additional visit will be performed as soon as possible, including the appropriate assessments that were specified in the trial schedule for the visit that was missed.

### Weekly monitoring calls

For intervention fidelity, the research assistants will be responsible to telephonically call the recruited patients to make sure that they are receiving and reading the messages, or getting the messages read to them, daily. Monitoring calls will be weekly for the first two months, where most of the intervention will be delivered in this period, followed by bi-weekly or monthly calls. This will also build rapport between the patient and the research team which will help in successful follow-ups and outcome assessments.

### Trial data management

REDCap, a secure web platform for building and managing online databases and surveys, will be used for managing the trial data. The REDCap mobile app will be used to collect the required data for the case report form (CRF) designed in the app. Screening forms will be programmed to collect data for differentiating eligible and non-eligible patients. At the screening stage, participants who meet all inclusion criteria and do not meet any exclusion criteria will be eligible to take part in the study. They will be given the participant information sheet and consent form. If they consent to take part in the study, their demographic details (including active mobile number) will be recorded on the REDCap database.

The mTB-Tobacco app will be linked to the REDCap server through API (Application Programming Interface) which will capture the patient trial ID along with the mobile numbers to send SMS text messages as per the defined schedule. The eligible and consented participants belonging to the intervention arm will receive the SMS messages, whereas those belonging to the control arm will not receive any SMS. Figure 3 gives a flowchart of data management procedures.

**Figure 3.**
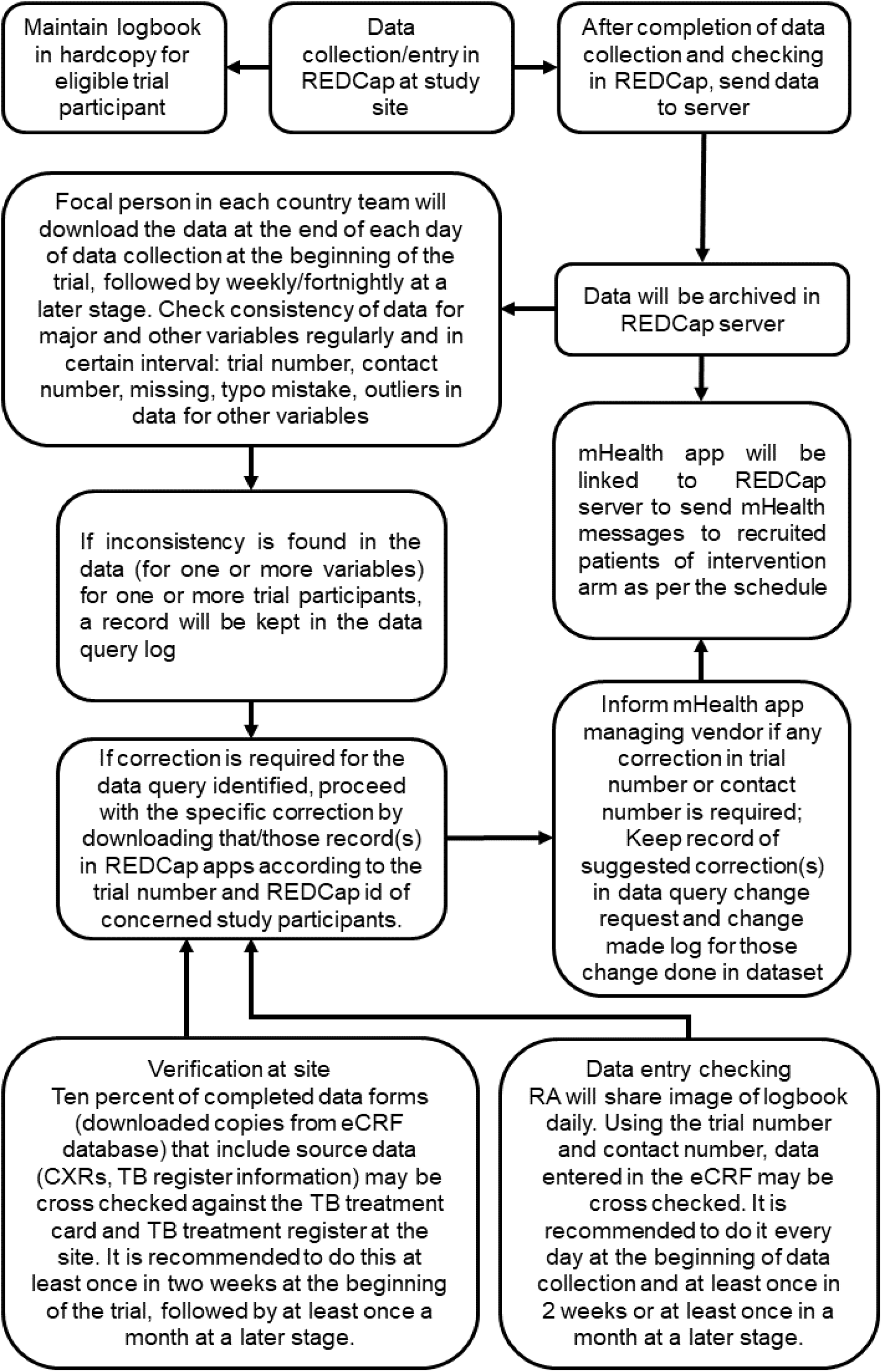
Flowchart of procedure of data management.

The data collection will consist of responses to a set of translated and structured questionnaires in the electronic Case Report Forms (CRF) which will be completed and facilitated by the field research team. The data entered will synchronise with the database on a secured server. De-identified data merged for study sites for separate eCRF and wide format data combining different eCRFs through matching and merging will be shared via Edinburgh DataSync with the trial statistician who will conduct the analysis.

Overall data quality will be ensured through training and supervision by the respective country trial team. Quantitative data once synced in the server will also be peer-reviewed by the statistician. The Trial Management Team and Country teams jointly will develop a detailed Data Management Plan to ensure data safety and quality.

### Sample size

The study requires a total of **2,716** smokers newly diagnosed with pulmonary TB (∼43 recruits from 63 TB centres, the clusters) assuming that 10% might not provide primary outcome data. This sample also includes the first 16 participants taking part in the pilot study, in case the intervention needed major refinement then we would be unable to use their data.

The required sample size for the superiority trial (phase 3) was determined by using a power of 90%, a significance level of 5%, and expected abstinence rates of 18% for mTB-Tobacco and 8% for normal treatment after 6 months.^19^ The study will comprise a total of 27 TB centres, considering the ratio of intervention to control as 2:1. After considering a 20% attrition rate, it was determined that the sample size would be 704 participants. This would result in an average of around 26 individuals in each cluster. Given that the study will be carried out as a cluster randomised control trial (cRCT) with an intracluster correlation coefficient (ICC) of 0.02,^20^ the design effect was computed to be 1.50. This results in an effective sample size of around **1080** people (or 40 subjects per site).

In the non-inferiority trial (phase 4), considering 90% power, one-sided hypothesis, and 2.5% significance level to establish non-inferiority of mTB-Tobacco intervention to face-to-face behavioural support, comparing 18 clusters in each treatment arm assuming face-to-face achieved 18% abstinence at six months and the non-inferiority margin was 8%. Assuming that just 2% would give up with no intervention at all, the established treatment effect of face-to-face behaviour support over natural cessation over 6 months would then be 16% (18%-2%). The non-inferiority margin of 8% equates to the mTB-Tobacco intervention preserving at least 50% (8/16) of the established treatment effect of the face-to-face intervention. The above-mentioned sample size assumptions will be checked mid-way through the recruitment period and the sample size will be adjusted accordingly. Considering a 20% attrition rate, the final sample size was determined to be 1036 participants, resulting in approximately 29 participants per cluster across 36 clusters (18 in each arm). The design effect was calculated to be 1.56, yielding an effective sample size of approximately **1620** subjects (or 45 subjects for each site). Further adjustments to the sample size assumptions will be made midway through the recruitment period as needed. Sample size details for superiority and non-inferiority trials are given as Supplementary Figures 4a and 4b respectively.

### Statistical analysis

#### Descriptive Statistics

Continuous outcomes will be summarised using means and standard deviations, and categorical outcomes using frequencies and percentages across intervention and control arms.

#### Inferential Statistics

A mixed-effects model will be used to assess the impact of mTB-Tobacco on continuous outcomes, incorporating fixed effects for the intervention, random effects for clusters, and relevant covariates. Logistic regression within the mixed-effects framework will be used to analyse categorical outcomes, accounting for cluster-level variability and intervention effects. for the analysis of data in this Cluster Randomised Controlled Trial. At the end of phase 3 superiority trial, an interim analysis will be done by an independent statistician to assess effectiveness of mTB-Tobacco intervention, the result will decide whether the trial needs to proceed to phase 4 non-inferiority trial or be terminated early, In this study, the missing data patterns will first be examined to determine whether the missingness adheres to patterns of complete randomness (MCAR), randomness (MAR), or non-randomness (MNAR). Multiple imputation, a robust technique that generates multiple datasets with imputed values for missing observations, will be utilised to address the missing data. As a sensitivity analysis technique, missing data imputation will be used to evaluate the robustness of our findings.^21^ The estimates of the intervention effect will next be compared between analyses carried out with complete case analysis and analyses utilising imputed datasets. This will improve the validity of the study’s conclusions by enabling an assessment of the effect of missing data on the estimation of intervention effects in this cluster-randomised controlled trial. JASP and R will be employed as our primary tools for data analysis.

#### Cost-effectiveness analysis

Health economic evaluation will be conducted to establish the value for money afforded by the mTB-Tobacco intervention. The cost of providing the interventions will be recorded and calculated using local cost data. Costs include staff, premises and pharmaceutical products. Patients’ wider use of public and private health care will also be recorded, using a self-report service use questionnaire. Patient cost profiles will be calculated by applying country-specific unit cost tariffs to quantities. Total intervention cost, costs of wider health care use and also patients’ out-of-pocket expenditure will be derived and a total cost will be applied to each patient in each trial arm.

In the superiority trial (phase 3), an incremental cost-effectiveness analysis (CEA) of the mTB-Tobacco intervention will be conducted, over and above usual care. EQ-5D-5L will be used to calculate Quality Adjusted Life Years (QALYs) using the area under the curve.^22^ Incremental cost-effectiveness ratios will be presented in terms of i) cost per QALY and ii) cost per additional quitter as based on the secondary outcome of the trial. A 12-month time horizon is selected for the CEA to utilise the longer-term window of healthcare use and changes in Health-Related Quality of Life (HRQoL). A full sensitivity analysis will be presented to assess the robustness of the results and the probability that mTB-Tobacco intervention is cost-effective at a range of threshold values.

In a non-inferiority trial (phase 4) resource quantities and attribute unit costs will be recorded for the mTB-Tobacco intervention and face-to-face behaviour support, inclusive of wider health system costs following the methodology of phase 3 superiority trial. Costs for each arm will be calculated and compared to a pre-defined acceptable cost difference. A cost-effectiveness analysis will also be done for the non-inferiority trial (phase 4). Differences in costs and QALYs between treatment arms will be estimated to calculate the incremental cost-effectiveness ratio (ICER) for the cost per QALY and cost per additional quitter.

Underlying uncertainty around the decision to adopt the intervention will be assessed using non-parametric bootstrap re-sampling technique. Bootstrapping is an efficient method for calculating the confidence limits for the ICER as its validity does not depend on any specific form of underlying distribution. We will perform the bootstrap 5,000 replications and construct the 95% confidence intervals for the ICERs based on the bootstrapping results. Cost-effectiveness acceptability curves (CEACs)^23^ will be constructed based on the bootstrap iterations to estimate the probability that the intervention is cost-effective at different threshold values for one QALY.

The ICER, calculated in terms of the cost per QALY, will be used to assess the value of money afforded by the intervention over and above the control and draw conclusions with respect to the potential cost-effectiveness of the intervention. To assess the impact of imputing missing data, we will also conduct a complete case analysis based on the participants who have both complete costs and QALYs at all time points, following the same analysis method as the primary analysis above. Sensitivity analyses using pattern mixture modelling will be used to examine the assumptions for multiple imputation methods.

#### User experience

Intervention fidelity and receipt of the SMS and face-to-face messages included in mTB-Tobacco will be assessed through a brief process evaluation using routinely collected data. Three questions in the Case Report Forms at week 9 follow up address:

a. whether or not any messages were received
b. the frequency of messages received and
c. the content of received messages (educational focus, medical focus, invitation to follow up, etc.)

The mTB-Tobacco fidelity data will be collected directly from the digital platform delivering messages. Data will be analysed for frequencies of reported answers, followed by regression analyses to assess:

1. Whether receiving any SMS is associated with quitting TB
2. Which frequency of receiving SMS is associated with a greater likelihood of quitting TB
3. Which type of information is associated with the greater likelihood of quitting TB

### Trial Steering Committee

An independent steering and monitoring committee will be set up to undertake the roles traditionally undertaken by the Trial Steering Committee (TSC) and Data Monitoring and Ethics Committee (DMEC). This committee will comprise independent members including a Chair and two other independent members. The independent members of the committee will be allowed to see unblinded data, but this data will not be reported to the other members of the research team. The responsibility of TSC would be to assist project managers in ensuring that the project is aligned with the objectives, manage risks, maintain project quality and track progress along with proposed timeframes. The committee will meet at least annually or more often as appropriate.

### Confidentiality and data protection

All laboratory specimens, evaluation forms, reports, and other records will be handled in a manner to maintain participant confidentiality. All records will be kept in a secure storage area with limited access. The Principal Investigator and study site staff involved in this study will not disclose or use for any purpose other than the performance of the study, any data, record, or other unpublished information, which is confidential or identifiable.

The research teams will comply with the requirements of data protection legislation (including the European Union General Data Protection Regulation, the Data Protection Act 2018 in the UK and any relevant Data Protection laws in Bangladesh and Pakistan, respectively with regard to the collection, storage, processing, and disclosure of personal information.

### Ethics and dissemination

Ethics approval was obtained from Edinburgh Medical School Research Ethics Committee, University of Edinburgh (EMREC), United Kingdom. Local ethics approval was obtained from Bangladesh Medical Research Council and the National Bioethics Committee for Research, Pakistan National Institutes of Health. The trial is registered on the ISRCTN registry: https://www.isrctn.com/ISRCTN86971818. Protocol amendments were also submitted and approved by all ethics committees. Separate manuscripts with the results of the superiority and non-inferiority phases will be submitted for publication in peer-reviewed journals, where authorship will be decided as per ICMJE criteria. On completion of the trial, de-identified data requests will be deposited in Edinburgh DataShare or DataVault, which are data repositories hosted by the University of Edinburgh. The data will be available to researchers upon reasonable request.

### Trial status

The latest version of the protocol (version 5.0, dated 25^th^ Jan 2024) was approved by EMREC, and local national ethics committees of Pakistan and Bangladesh. This paper is a structured version of currently approved protocol, complying with SPIRIT guidelines to report protocols of randomised controlled trials.^24^ Recruitment of participants was started in February 2024 for the superiority trial in Pakistan and Bangladesh, where the recruitment completion date is anticipated to be within the next 6 months for phase 3. The results of phase 3 will be analysed after 6-month follow-up of the last recruited patient, depending on the analysis of the results the principal investigator will decide whether to move to the non-inferiority trial phase or to end the trial early. The total duration of this study is three years (Jun 2022 - June 2025).

## DISCUSSION

This trial will be the first one to test the effectiveness of mHealth based mTB-Tobacco intervention to help TB patients quit smoking at 6 months. The sample size is appropriately calculated keeping in mind the cluster design and design effect to appropriately power this trial to test the primary objective. The mHealth based interventions are promoted by policymakers due to low resource requirements. An effective mTB-Tobacco intervention could be transformational for TB patients who smoke. It will not only benefit the TB patients, but will also allow the national TB control programmes to have a less resource-intensive and effective intervention for smoking cessation that can be easily integrated in their system. A shift from paper based-record TB recording and reporting system to a digital system in Pakistan and Bangladesh makes it easier to integrate mHealth intervention into the existing system.

To date, all evidence-based smoking cessation interventions, face-to-face behaviour interventions, nicotine replacement therapies and pharmacotherapy-based interventions have failed to be integrated into existing TB control programmes. A case study was published where challenges of integrating tobacco cessation interventions in TB programs were explored in the context of Pakistan and Nepal.^25^ The main challenges that were highlighted in the study included health professionals’ doubts about the contextual relevance of interventions, non-conducive environments, political reluctance to change, workload of TB staff, motivation of TB staff and capacity to deliver cessation intervention and inadequate training and support during intervention implementation.^25^ Several of the above-mentioned issues can be resolved through a digitally delivered intervention. For instance, the TB staff would not have to perform any extra task out of their routine to deliver the intervention as the SMS messages are delivered to the TB patient’s personal mobile phone number through an automated system. In future, the digital app can be further improved by making it interactive instead of just one-way delivery. It can also be personalised for every TB patient who wants to quit smoking by using artificial intelligence tools. The cost of the intervention will be one-off, for example, the web portal that will be developed under this project can be used by TB programmes with no to minimal extra cost to only account for the SMS text message delivery.

Keeping in mind that TB disease is more prevalent in vulnerable populations e.g. lower socioeconomic groups,^26^ and such population is more likely to have poor literacy as compared to the general population. This may be a challenge as TB patients’ ability to read and understand SMS messages may be a barrier. In this trial, we will take help from a literate family member in such instances. Future studies may test recorded voice messages in the local language. Overall literacy and digital literacy are improving in LMIC which means that there will be literate, tech-savvy societies in the future.

In conclusion, this study may lead to a substantial impact if mTB-Tobacco intervention enhances quitting smoking among TB patients, leading to better treatment outcomes and reducing the disease and economic burden of TB and tobacco. The intervention can be scaled up and contribute towards achieving the Sustainable Development Goals (SDG 3.0 - Improving health and well-being, SDG 3.3 - Fighting communicable diseases and SDG 3.A - Implement the WHO Framework Convention on Tobacco Control).

## Supporting information

Spirit checklist

all suppl material

## Data Availability

All data produced in the present study are available upon reasonable request to the authors

## Funding

This study is funded by the National Institute of Health Research UK under a research award named NIHR Global Health Research Unit on Respiratory Health (RESPIRE) (Award ID: NIHR132826) where the contracting organisation is The University of Edinburgh, while implementing partners include The Initiative (Pakistan) and ARK Foundation (Bangladesh). The funders had no role in the design of the study, data collection, analysis, interpretation or in writing the manuscript

## Disclaimer

The views expressed in this publication are those of the author(s) and not necessarily those of the NIHR or the UK Government. The RESPIRE collaboration comprises the UK and LMIC Grant holders, Partners and research teams as listed on the RESPIRE website (www.ed.ac.uk/usher/respire).

## Acknowledgement

The authors thank the National TB Programs of Pakistan and Bangladesh for facilitating the study. This research was funded by the UK National Institute for Health and Care Research (NIHR) (Global Health Research Unit on Respiratory Health (RESPIRE); NIHR132826) using UK aid from the UK Government to support global health research.

## Collaborator/Sponsor

NIHR Global Health Research Unit on Respiratory Health (RESPIRE) at the University of Edinburgh, UK (www.ed.ac.uk/usher/respire). The Sponsor will assess the study to determine if an independent risk assessment is required. If required, the independent risk assessment will be carried out by the ACCORD Quality Assurance Group to determine if an audit should be performed before/during/after the study and, if so, at what frequency.

## Contributors

MZ compiled the manuscript for publication, and contributed in introduction and discussion writing; FR, AC and SHR contributed to developing the data management section; AKL contributed in manuscript compilation and write-up; SA contributed in the methodology section of mHealth intervention and provided technical input regarding digital appl MD contributed in statistical analysis and sample size calculation section writing; MB contributed in user experience survey section; SP contributed in economic analysis section, AK (PI from Pakistan) and RH (PI from Bangladesh) contributed in manuscript writing, proof-reading and supervision of local research teams, KS (chief PI) contributed in research idea conception, acquiring funding, manuscript proof-reading and overall supervision

## Conflict of interest

There is no conflict of interest to be declared by any author.

## Notes

### Competing Interest Statement

The authors have declared no competing interest.

### Clinical Trial

ISRCTN86971818

### Author Declarations

National Bioethics Committee gave ethical approval for this work

