## Supplementary material for "A mHealth intervention (mTB-Tobacco) for smoking cessation in people with drug-sensitive pulmonary tuberculosis: protocol for an adaptive design, cluster randomised controlled trial (Quit4TB)": all suppl material

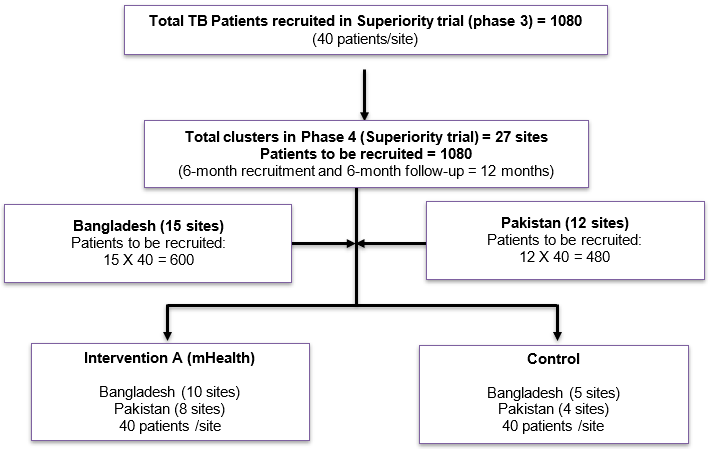

**Supplementary Figure 1a: Cluster numbers and cluster sizes for the superiority trial in Pakistan and Bangladesh**

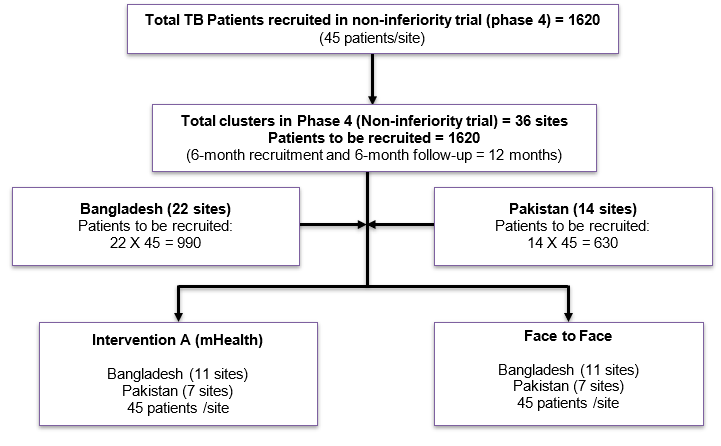

**Supplementary Figure 1b: Cluster numbers and cluster sizes for the non-inferiority trial in Pakistan and Bangladesh**

**Supplementary Table 1: List of text messages included in mTB-Tobacco intervention along with delivery schedule**

| **Day Received** | **KEYWORD CATEGORY** | **Message** | **Character Count** |
| --- | --- | --- | --- |
| Day Registered  Q (-8)^[1]^  Tx (+1)^[2]^ | Generic  (for all) | 1. Welcome to mTB-Tobacco! Congratulations on deciding to QUIT tobacco use and improve your chances of successfully recovering from TB.   Time: On day of enrolment | 133 |
|  | Generic  (for all) | 2. Well done for joining this programme. We will help you quit tobacco and adhere to TB medication by giving useful tips throughout the treatment course.   Time: On day of enrolment | 151 |
|  | Generic  (for all) | 3. You will get up to 5 text messages every day to help you successfully recover from TB. All messages received in mTB-Tobacco will be free.   Time: On day of enrolment | 146 |
| **mTB-Tobacco intervention phase!** | | | |
| Q (-7)  Tx (+2) | Generic  (for all) | 4. TB germs spread in the air when TB patient coughs, sneezes or spits. TB is curable! You must take the medicine regularly for full [*xx*] months as your doctor tells.   Time: 9:00 am in morning | 163 |
|  | Generic  (for all) | 5. Deciding to quit tobacco for good is a great step. You should feel proud of yourself. We’ll send you tips to help you get ready!   Time: 12:00 pm | 128 |
|  | Generic  (for all) | 6. The best way to quit tobacco use is to stop completely. To help you quit, we have set a date for you, 7-days from today, to stop using tobacco completely.   Time: 5:00 pm | 159 |
|  | Generic  (for all) | 7. Remember, it will **NOT** be helpful if you:  - slowly decrease tobacco use - use tobacco once in a while - or replace it with other kinds of tobacco such as [*waterpipe, bidi, smokeless tobacco*]   Time: 7:00 pm | 159 |
| Q (-6)  Tx (+3) | Generic  (for all) | 8. Do you know that tobacco use can stop your recovery from TB? People who do not quit are more likely to have heart disease and cancer.   Time: 9:00 am | 139 |
|  | Generic  (for all) | 9. Quitting tobacco will help with your recovery!   The benefits of quitting will start within days and will continue during and after treatment.  Time: 12:00 pm | 140 |
|  | Generic  (for all) | 10. Within 1 month, your cough and sputum production will decrease and you will be able to breathe easily. If you don’t quit, your recovery will be delayed**۔**   Time: 2:00 pm | 151 |
|  | Generic  (for all) | 11. Within 6 months of quitting, you will be cured of TB as long as you take your medication.   Don’t forget to take your TB medication regularly!  Time: 5:00 pm | 139 |
|  | (smoking forms only) | 12. If you don’t quit you will have a higher chance of getting TB again! Craving to smoke is temporary but damage to your lungs is permanent!   Time: 7:00 pm | 137 |
|  | ^[3]^(chewing tobacco) | 13. If you don’t quit you will have a higher chance of getting TB again!   Tobacco contains harmful substances and chewing can lead to mouth cancer!  Time: **8**:00 pm | 147 |
| Q (-5)  Tx (+4) | Generic  (for all) | 14. Tell your family and friends that you have TB so they can offer you support and help you to get better. TB does not spread by shaking hands, cuddling or hugging.   Time: 9:00 am | 161 |
|  | Generic  (for all) | 15. Remember that your QUIT Date is near! 5 days to go! Share this date with your family/friends for support to stay tobacco free and continue taking TB medicine.   Time: 12:00 pm | 160 |
|  | Generic  (for all) | 16. It is important to get your family and friends support during treatment. TB is spread through the air, so you will not transmit TB by sharing food or utensils.   Time: 02:00 pm | 160 |
|  | Generic  (for all) | 17. Remember, there is **NO** safe or mild tobacco. Bidi, cigarette, hookah and chewing tobacco are all harmful. (Local tobacco product) is also harmful.   Time: 05:00 pm | 145 |
| Q (-4)  Tx (+5) | Generic  (for all) | 18. It takes 6 months of regular treatment to kill TB germs but after few weeks of taking medicine you can continue life as normal. Take your TB medication regularly   Time: 9:00 am | 163 |
|  | Generic  (for all) | 19. 12 months after quitting your body will be highly resistant to catch TB again!   Time: 12:00 pm | 78 |
|  | Generic  (for all) | 20. PREPARE TO GET RID OF TOBACCO PRODUCTS; matches, ashtrays, paan dan/chewing box, water-pipe, anything that might remind you of tobacco use.   Time: 02:00 pm | 141 |
|  | Generic  (for all) | 21. Make a list of your reasons for quitting tobacco. Health, family, and saving money are good reasons to qui   Time: 05:00 pm | 111 |
|  | (smoking forms only) | 22. Did you know that smoking can cause sexual problems? It can even cause impotence.   Time: 07:00 pm | 81 |
|  | (chewing tobacco) | 23. Did you know that chewing can cause staining of the teeth, bad breath and gum problems? It can even cause throat cancer.   Time: 08:00 pm | 120 |
| Q (-3)  Tx (+6) | Generic  (for all) | 24. While preparing for your quit date, do not forget your TB medicines! Do not miss even a single dose, it will take longer to cure.   Time: 9:00 am | 129 |
|  | Generic  (for all) | 25. When you first start using tobacco your brain changes and expects regular doses of nicotine. When you quit, nicotine is not available and you feel an urge for it.   Time: 12:00 pm | 162 |
|  | Generic  (for all) | 26. The urge for nicotine can be very strong and can undermine your motivation to quit, especially on occasions and situations that remind you of tobacco.   Time: 02:00 pm | 150 |
|  | Generic  (for all) | 27. You are moving towards your QUIT date! 3 days to go! It is important that you request family and friends who use tobacco about not using it when they are with you.   Time: 05:00 pm | 163 |
|  | Generic  (for all) | 28. Before your QUIT date, please think through things that tempt your desire to use tobacco, like certain times, or places or people. Write these situations down.   Time: 07:00 pm | 160 |
| Q (-2)  Tx (+7) | Generic  (for all) | 29. Tobacco use will make it harder to cure your TB. Quitting will help you recover faster from TB as long as you take your medication.   Time: 9:00 am | 131 |
|  | Generic  (for all) | 30. Did you think about situations that might trigger your desire to use tobacco? Start planning for how you will manage your urges to use tobacco.   Time: 12:00 pm | 144 |
|  | Generic  (for all) | 31. To avoid the urges to use tobacco, think of alternative things you could do instead, like using gum, clove or cardamom to keep your mouth busy.   Time: 02:00 pm | 143 |
|  | Generic  (for all) | 32. Many people use tobacco to deal with stress, tension or boredom. Try walking, doing exercise, yoga, deep breathing, meditation or prayer instead.   Time: 05:00 pm | 145 |
|  | Generic  (for all) | 33. It is normal to be nervous about quitting. Don’t panic, your family and friends are there to support you! Tell them that you are ready to quit for good!   Time: 07:00 pm | 152 |
| Q (-1)  Tx (+8) | Generic  (for all) | 34. Remember, TB is curable if you take the advised treatment. However, outcome is poorer in tobacco users. Quitting will help you successfully recover from TB.   Time: 09:00 am | 157 |
|  | Generic  (for all) | 35. Within first few hours of stopping tobacco use, your brain will start getting used to being without nicotine. This adjustment may result in craving or withdrawal.   Time: 12:00 pm | 162 |
|  | Generic  (for all) | 36. Cravings can be uncomfortable and you might feel uneasy. Rest assured that these are perfectly normal and will reduce overtime as you progress with your quit.   Time: 02:00 pm | 158 |
|  | Generic  (for all) | 37. After quitting you may feel constipated, irritable, restless or lightheaded due to withdrawal from nicotine that you may or may not get. These will go away soon! Consult with your TB Provider if needed.   Time: 05:00 pm | 161 |
|  | Generic  (for all) | 38. Ready for quitting tomorrow! Hope you have removed all tobacco products, matches, ashtrays, paan dans, and waterpipe around you. If not do so right away!   Time: 07:00 pm | 153 |
| QUIT DATE (Q) This is the 8th day since joining mTB-Tobacco | Generic  (for all) | 39. You need clean air to fight against TB germs. Ask your family and friends NOT to smoke around you. Continue taking your TB medicines!   Time: 09:00 am | 133 |
|  | Generic  (for all) | 40. Today is your QUIT DATE! Remove all tobacco products around you. You can do it!   Time: 12:00 pm | 78 |
|  | Generic  (for all) | 41. Drink lot of fluids (water, coconut water, lime juice, lassi, buttermilk) today to feel fresh and fight cravings. Avoid sugary drinks or alcohol.   Time: 02:00 pm | 143 |
|  | Generic  (for all) | 42. Have gum/cloves/cardamom/mints, water, and healthy snacks to get you through the day. You have made the right choice!   Time: 05:00 pm | 118 |
|  | Generic  (for all) | 43. Tell your near and dear ones that you are trying to quit tobacco today. Ask for their support to stay tobacco free.   Time: 07:00 pm | 115 |
|  |  | 44. If you’re feeling stressed or upset before sleeping at night, do not smoke tobacco. Tobacco use never solved any problem for you. You did it. THINK POSITIVE.   Time: 09:00 pm |  |
| Q (+1)  Tx (+10) | Generic  (for all) | 45. 24 hours tobacco free …well done! Be sure to reward yourself. Celebrate with your family and friends.   Time: 09:00 am | 101 |
|  | Generic  (for all) | 46. The first days are going to be tough. Be patient. Don’t switch from one kind of tobacco product to another. Do not use (local tobacco product).   Time: 05:00 pm | 143 |
|  | Generic  (for all) | 47. Remind yourself of your reasons for quitting. That will help keep you on track when you need a boost.   Time: 07:00 pm | 101 |
| Q (+2)  Tx (+11) | Generic  (for all) | 48. To be cured from TB you must remember to keep taking your medicines but having a healthy lifestyle will also help you recover better.   Time: 09:00 am | 135 |
|  | Generic  (for all) | 49. It is important to eat healthy snacks (e.g., peanuts, raisins, aniseed) with proteins and vitamins frequently and in ample quantities. Make sure you get plenty of rest!   Time: 12:00 pm | 135 |
|  | Generic  (for all) | 50. Refrain from consuming alcohol. Breathe clean fresh air and keep it tobacco free. Ask family and friends not to smoke indoors!   Time: 02:00 pm | 126 |
|  | Generic  (for all) | 51. Refrain from using any kind of tobacco. Bidi, cigarette, hookah and chewing tobacco are all harmful. (Local tobacco product) is also harmful.   Time: 05:00 pm | 141 |
|  | Generic  (for all) | 52. If your urge to smoke is because you’re feeling stressed, try closing your eyes & taking deep breaths. Tell yourself that smoking is a false way of coping!   Time: 07:00 pm | 155 |
| Q (+3)  Tx (+12) | Generic  (for all) | 53. 3rd day without tobacco! Your food will taste better. Drink plenty of water to avoid constipation. You may begin to feel better. Keep taking your TB medicine!   Time: 12:00 pm | 158 |
|  | Generic  (for all) | 54. Talk to someone who has already QUIT or spend time with persons who do not use tobacco.   Time: 02:00 pm | 87 |
|  | Generic  (for all) | 55. Keep your mouth busy! Use chewing gum, cloves, cardamom, or mints to avoid tobacco use.   Time: 05:00 pm | 87 |
| Q (+4)  Tx (+13) | Generic  (for all) | 56. 4 days tobacco free! The first few days can be tough. Try to relax. Listen to music, have fresh fruits, do yoga or deep breathing.   Time: 12:00 pm | 130 |
|  | Generic  (for all) | 57. Soon after starting your TB medicines, you may begin to feel better but you are not completely cured. You must keep taking your medicine!   Time: 02:00 pm | 138 |
|  | Generic  (for all) | 58. Sometimes the TB medicine might have side-effects that make you want to stop taking it. Be patient, symptoms will subside. Just keep taking the medicines.   Time: 05:00 pm | 154 |
| Q (+5)  Tx (+14) | Generic  (for all) | 59. Well done for staying tobacco free and regularly taking your TB medicine. Your body is already starting to heal. Keep up the good work.   Time: 09:00 am | 136 |
|  | Generic  (for all) | 60. Day 5! Stay positive. Do not let things get you down. The journey towards a tobacco and TB free life is definitely worth it.   Time: 12:00 pm | 124 |
|  | Generic  (for all) | 61. Drink lots of fluids like lime juice, buttermilk, coconut water. Avoid cold drinks and alcohol.   Time: 02:00 pm | 95 |
|  | Generic  (for all) | 62. Stay away from people and places that remind you of tobacco. Do things which help you to relax. Some people find gardening, sewing, or watching a movie helpful.   Time: 05:00 pm | 160 |
| Q (+6)  Tx (+15) | Generic  (for all) | 63. Get your family and friends support as your body combats TB and you deal with cravings. Ask a family member or friend to remind you to take your medicines.   Time: 09:00 am | 155 |
|  | Generic  (for all) | 64. During festivals it is easy to forget taking medicines. Ask your friends and family members to remind you. Always carry your medicines while traveling!   Time: 12:00 pm | 153 |
|  | Generic  (for all) | 65. 6 days tobacco free! Your body is free from tobacco related toxins. Stay active or go for a walk; it helps to reduce stress.   Time: 02:00 pm | 126 |
|  | Generic  (for all) | 66. If you’re feeling stressed or upset, talk to someone close to you for support.   Time: 05:00 pm | 78 |
| Q (+7)  Tx (+16) | Generic  (for all) | 67. 1 week tobacco free! Do not look back now. Feel happy and do something special today to celebrate your success!Time: 09:00 am | 111 |
|  | Generic  (for all) | 68. Find out how much money you have saved from not using tobacco in this week. Think about what else you can do with the saved money!   Time: 12:00 pm | 131 |
|  | Generic  (for all) | 69. If you have the urge to use tobacco, watch a movie, go out or buy yourself something as reward for staying QUIT.   Time: 05:00 pm | 112 |
| Q (+8)  Tx (+17) | Generic  (for all) | 70. Remember to keep coming to see the TB provider for your appointments or if you are having any problems. You will recover from TB if you keep taking your medicine.   Time: 09:00 am | 162 |
|  | Generic  (for all) | 71. Use a calendar to check off the days you have taken your TB medicines and the days that you have stayed tobacco free!   Time: 05:00 pm | 117 |
| Q (+9)  Tx (+18) | Generic  (for all) | 72. Quitting tobacco makes you feel healthier and energetic.   Time: 09:00 am | 55 |
|  | Generic  (for all) | 73. Tobacco smoke is dangerous for everyone, including non-smokers who inhale this smoke. Avoid places where others are smoking.   Time: 12:00 pm | 124 |
|  | Generic  (for all) | 74. Smoking spreads TB germs. Make your home tobacco free!   Time: 05:00 pm | 54 |
| Q (+10)  Tx (+19) | Generic  (for all) | 75. Hope you are feeling good! Keep up that positive attitude and stay strong. Stay away from all tobacco products, even those prepared locally.   Time: 09:00 am | 141 |
|  | Generic  (for all) | 76. Tobacco use does not make anything better. Stay strong and say NO if anyone asks you to smoke/chew tobacco. Be proud that you said NO.   Time: 05:00 pm | 134 |
| Q (+11)  Tx (+20) | Generic  (for all) | 77. Eat nutritious and healthy food. It will help you stay well and get better faster.   Remember to keep taking your TB medicine!  Time: 09:00 am | 124 |
|  | Generic  (for all) | 78. Avoid fried food when you have craving. Eat vegetable, salad, fruit, drink lassi, milk, coconut water.   Time: 05:00 pm | 150 |
| Q (+12)  Tx (+21) | Generic  (for all) | 79. Exercise regularly, it helps to strengthen your body defences against TB germs.   Time: 09:00 am | 79 |
|  | Generic  (for all) | 80. Gaining a little weight after quitting may occur for some time, but healthy eating and exercise can prevent this.   Time: 05:00 pm | 113 |
| Q (+13)  Tx (+22) | Generic  (for all) | 81. Remember to get more medicines you run out! Check your next appointment date with TB provider and mark the calendar!   Time: 09:00 am | 123 |
|  | Generic  (for all) | 82. Stay positive by exercising, doing yoga, meditation or starting a new hobby. Make friends with people who do not use tobacco!   Time: 02:00 pm | 125 |
|  | Generic  (for all) | 83. Adopt a healthy lifestyle. Reduce your chances of ever developing TB again.   Time: 05:00 pm | 75 |
| Q (+14)  Tx (+23) | Generic  (for all) | 84. Well done, 2 weeks tobacco free! How much time, energy, health, and money you have saved! You deserve a treat-make today special!   Time: 09:00 am | 129 |
|  | Generic  (for all) | 85. Nobody likes a dirty mouth and bad breath. Now your smile is brighter and your mouth healthier.   Time: 02:00 pm | 95 |
|  | Generic  (for all) | 86. Remind people close to you that you have QUIT tobacco. They want only what is best for you and that means staying QUIT and healthy.   Time: 05:00 pm | 131 |
| Q (+15)  Tx (+24) | Generic  (for all) | 87. Congratulations! Being tobacco free means you are not a slave to something that actually harms you.   Time: 09:00 am | 99 |
| Q (+16)  Tx (+25) | Generic  (for all) | 88. Well done for taking your TB medicines regularly. You must be feeling much better now. Your next monthly follow-up appointment is in 5 days!   Time: 09:00 am | 141 |
|  | Generic  (for all) | 89. 16 days! Tobacco use never solved any problem for you. YOU DID IT.   You can do great things, so think positively.  Time: 12:00 pm | 111 |
| Q (+17)  Tx (+26) | Generic  (for all) | 90. Share your TB experience with others and motivate them to seek help if needed! Ask your near ones to quit tobacco use as it increases their risk of getting TB.   Time: 09:00 am | 159 |
|  | Generic  (for all) | 91. Keep up the great effort! Spend time with someone close to you. Go to places where no one smokes or uses tobacco.   Time: 02:00 pm | 113 |
|  | Generic  (for all) | 92. Avoid people or places that remind you of using tobacco or going near shops that sell tobacco. It will make coping easier.   Time: 05:00 pm | 122 |
| Q (+18)  Tx (+27) | Generic  (for all) | 93. Your next monthly follow-up appointment is in 3 days! Do not forget to visit your TB provider and collect more medicine.   Time: 09:00 am | 120 |
|  | Generic  (for all) | 94. If you are bored, find something to do, so you won’t feel tempted to use tobacco. Plan an outing, spend time with friends and family.   Time: 02:00 pm | 133 |
|  | Generic  (for all) | 95. You used to pay money for something that would kill you. Be happy you are not a target anymore.   Time: 05:00 pm | 95 |
| Q (+19)  Tx (+28) | Generic  (for all) | 96. Encourage your friends and family to get tested for TB, if they live with a TB patient. Remember, TB spreads through the air.   Time: 09:00 am | 125 |
|  | Generic  (for all) | 97. Remember, change is POSSIBLE and for your good. Do not give up your resolve to QUIT tobacco. Even if you fall, get up and try again.   Time: 05:00 pm | 132 |
| Q (+20)  Tx (+29) | Generic  (for all) | 98. Lung capacity increases by 30% after a few weeks without smoking! Breathe deeply and take a walk.   Time: 09:00 am | 96 |
|  | Generic  (for all) | 99. Distract yourself during a craving by being active. Physical activity can help you stay QUIT and improves your mood.   Time: 05:00 pm | 116 |
| Q (+21)  Tx (+30) | Generic  (for all) | 100. 3 weeks tobacco free! Reward yourself again. Go for a walk with someone close, buy a small gift or cook something nice.   Time: 09:00 am | 119 |
|  | Generic  (for all) | 101. And well done for successfully completing 1 month of TB treatment! Your monthly follow-up appointment is today, please do not forget to go!   Time: 09:30 am | 139 |
| Q (+22)  Tx (+31) | Generic  (for all) | 102. Be a role model; motivate your friends and family members to quit tobacco use and get tested for TB if they live with a TB patient.   Time: 09:00 am | 131 |
|  | Generic  (for all) | 103. Don’t be fooled by tobacco and (local tobacco product) advertisements and promotion. You now know the REAL truth about smoking & other tobacco use. Tell others.   Time: 05:00 pm | 160 |
| Q (+23)  Tx (+32) | Generic  (for all) | 104. Hope you collected more medicine for your TB treatment. Write yourself a note and put it on the fridge or a door. Keep taking medicines and stay healthy!   Time: 12:00 pm | 153 |
|  | Generic  (for all) | 105. The majority of tobacco users wish they had never started. Be happy with your decision to QUIT tobacco.   Time: 02:00 pm | 103 |
|  | Generic  (for all) | 106. All forms of tobacco are harmful. Remember, tobacco use causes high blood pressure, heart attack, TB, paralysis and cancers.   Time: 05:00 pm | 124 |
| Q (+24)  Tx (+33) | Generic  (for all) | 107. Continue taking your TB medicines! In case, you missed your dose, take the next dose as scheduled.   Time: 09:00 am | 97 |
|  | Generic  (for all) | 108. Quitting tobacco not only helps your lungs heal, save you from cancer but makes you healthier and stronger.   Time: 05:00 pm | 108 |
| Q (+25)  Tx (+34) | Generic  (for all) | 109. Ask a person close to you how your breath is after Quitting. Your breath is fresh! No bad breath! Aren’t you glad you QUIT?   Time: 02:00 pm | 123 |
| Q (+26)  Tx (+35) | Generic  (for all) | 110. Be careful when you go out. Do not let yourself slip and use tobacco again. You have come so far. Remember to take your TB medicine!   Time: 02:00 pm | 131 |
| Q (+27)  Tx (+36) | Generic  (for all) | 111. Don’t forget to reward yourself for staying tobacco free! Watch a movie, buy something, or put away the money you have saved in a money box.   Time: 02:00 pm | 140 |
| Q (+28)  Tx (+37) | Generic  (for all) | 112. Alcohol may increase your urge to use tobacco and cause a relapse. Stay away from alcohol, cold drinks and (local tobacco products).   Time: 02:00 pm | 132 |
|  | Generic  (for all) | 113. Alcohol also damages your liver and your body’s defence against infections such as TB and HIV. Cutting down on alcohol will help recover faster from TB.   Time: 05:00 pm | 152 |
| Q (+29)  Tx (+38) | Generic  (for all) | 114. Don’t let stress undo all your hard work. You CAN deal with it without tobacco. Go for walk, exercise, take deep breaths. Stay healthy, keep taking TB medicine!   Time: 02:00 pm | 159 |
| Q (+30)  Tx (+39) | Generic  (for all) | 115. Congrats! 1 MONTH since you became tobacco free! It is a major milestone. Share the good news with your family and friends. Do something special!   Time: 12:00 pm | 145 |
| Q (+31)  Tx (+40) | Generic  (for all) | 116. Tobacco users face an increased risk of sexual problems. Be glad you have decided to QUIT. Stay tobacco free!   Time: 12:00 pm | 109 |
| Q (+32)  Tx (+41) | Generic  (for all) | 117. If you stop taking TB medicines, you can pass TB germs to others! Keep taking your medicine to help you recover and protect others from TB!   Time: 12:00 pm | 139 |
| Q (+33)  Tx (+42) | Generic  (for all) | 118. BREATHE DEEPLY! Inhale through nose, exhale through mouth, repeat 10 times. This will help you get over the urge for tobacco.   Time: 12:00 pm | 125 |
| Q (+34)  Tx (+43) | Generic  (for all) | 119. All kinds of tobacco reduce life span and make you look aged. Smoking or chewing tobacco is NOT safe.   Time: 12:00 pm | 101 |
| Q (+35)  Tx (+44) | Generic  (for all) | 120. By taking TB medicines you can prevent TB disease and keep your family healthy! Don’t forget to take your medicines!   Time: 09:00 am | 116 |
|  | Generic  (for all) | 121. Even now, some things may make you want to use tobacco again! Use the skills you have learned to get through your craving without tobacco.   Time: 05:00 pm | 138 |
| Q (+36)  Tx (+45) | Generic  (for all) | 122. When faced with any difficulty, try to overcome it. Quitting tobacco is no different. Do not look back now. You have come a long way.   Time: 02:00 pm | 133 |
| Q (+37)  Tx (+46) | Generic  (for all) | 123. Congrats! It’s been 1.5 months since you started TB treatment. Continue your commitment with taking TB medicines regularly!   Time: 09:00 am | 123 |
|  | Generic  (for all) | 124. Think of what tobacco use costs you in one year. Think about all the other things you could have done with that money. Keep up the tobacco free lifestyle!   Time: 12:00 pm | 154 |
| Q (+38)  Tx (+47) | Generic  (for all) | 125. Arguments may still trigger desire for tobacco. To get relief from stress, practice deep breathing and meditation.   Time: 12:00 pm | 114 |
| Q (+39)  Tx (+48) | Generic  (for all) | 126. If you slip and use tobacco again, keep trying to QUIT. Learn from your mistakes this time.   Time: 12:00 pm | 91 |
| Q (+40)  Tx (+49) | Generic  (for all) | 127. Remember, if you stop taking your TB medicines or don’t take it correctly, your medicine may lose the power to cure your TB!   Time: 09:00 am | 125 |
|  | Generic  (for all) | 128. Pregnant women who use tobacco or are exposed to others’ tobacco smoke are at risk for stillbirths & low weight babies.   Time: 05:00 pm | 79 |
| Q (+41)  Tx (+50) | Generic  (for all) | 129. Remember to take your TB medicine in full dose, even when you don’t feel good. It is the only way to kill TB germs.   Time: 09:00 am | 115 |
|  | Generic  (for all) | 130. This is the beginning of a new life for you. Now you know the way to succeed. Stay tobacco free and help others in quitting tobacco!   Time: 02:00 pm | 132 |
|  | Generic  (for all) | 131. Now that you are tobacco free, encourage others to QUIT۔   Time: 05:00 pm | 56 |
| **mTB-Tobacco Follow-up phase!** | | | |
| Q (+52)  Tx (+61) | Generic  (for all) | 132. Well done! You have successfully completed first 2 months of your TB treatment. Now it’s time to visit your provider for sputum test and collect more medicine!   Time: 12:00 pm | 159 |
| Q (+112)  Tx (+121) | Generic  (for all) | 133. Well done! You have successfully completed 4 months of your TB treatment. Now it’s time to visit your provider for sputum test and collect more medicine!   Time: 12:00 pm | 153 |
| Q (+180)  Tx (+189) | Generic  (for all) | 134. Well done! You have successfully completed 6 months of your TB treatment. Now it’s time to visit your provider for the final sputum test.   Time: 12:00 pm | 137 |

1. This is number of days to the quit date
2. Tx is used for TB treatment and +1 means the first day of treatment
